# Tryptophan pathway metabotypes associate with disease activity and immune-metabolic dysfunction in inflammatory bowel disease

**DOI:** 10.64898/2026.05.03.26352309

**Authors:** Danielle M. M. Harris, Arno R. Bourgonje, Peder Rustøen Braadland, Cathy McShane, Lina Welz, Silvio Waschina, Susanne Ibing, Florian Tran, Bruce E. Sands, Marla Dubinsky, Mayte Suarez-Farinas, Carmen Argmann, Per M Ueland, Adrian McCann, Trond Espen Detlie, May-Bente Bengtson, Vendel Kristensen, Andre Franke, Jean-Frédéric Colombel, Philip Rosenstiel, Kenneth Croitoru, Harry Sokol, Williams Turpin, Johannes Roksund Hov, Marte Lie Høivik, Ryan C. Ungaro, Stefan Schreiber, Konrad Aden

**Affiliations:** Institute of Clinical Molecular Biology, University Medical Center Schleswig-Holstein and Christian-Albrechts-University of Kiel, Kiel, Germany; Department of Internal Medicine I, University Medical Center Schleswig-Holstein, Kiel, Germany; Institute for Human Nutrition and Food Science, Division of Nutriinformatics, Christian-Albrechts-University of Kiel, Kiel, Germany; The Henry D. Janowitz Division of Gastroenterology, Department of Medicine, Icahn School of Medicine at Mount Sinai, New York, NY, USA; Department of Gastroenterology and Hepatology, University Medical Center Groningen, University of Groningen, Groningen, the Netherlands; Research Institute of Internal Medicine, Oslo University Hospital, Oslo, Norway; Faculty of Medicine, Institute of Clinical Medicine, University of Oslo, Oslo, Norway; Norwegian PSC Research Center, Department of Transplantation Medicine, Oslo, University Hospital, Oslo, Norway; Lunenfeld-Tanenbaum Research Institute, Mount Sinai Hospital, Toronto, Ontario, Canada; Department of Genetics and Genomic Sciences, Icahn School of Medicine at Mount Sinai, New York, NY, USA; Hasso Plattner Institute, Digital Engineering Faculty, University of Potsdam, Potsdam, Germany; Department of Population Health Science and Policy, Icahn School of Medicine at Mount Sinai, New York, NY, USA; Bevital AS, Bergen, Norway; Department of Gastroenterology, Akershus University Hospital, Lørenskog, Norway; Department of Gastroenterology, Vestfold Hospital Trust, Tønsberg, Norway; Department of Gastroenterology, Oslo University Hospital, Oslo, Norway; Division of Gastroenterology and Hepatology, Temerty Faculty of Medicine, University of Toronto, Toronto, Ontario, Canada; Sorbonne Université, INSERM UMRS-938, Centre de Recherche Saint-Antoine, CRSA, AP-HP, F-75012 Paris, France; Gut, Liver & Microbiome Research (GLIMMER) FHU, Paris, France; Université Paris-Saclay, INRAe, AgroParisTech, Micalis Institute, Jouy-en-Josas, France

## Abstract

**Background:** Tryptophan (Trp) metabolism is a central immunometabolic axis in inflammatory bowel disease (IBD) and has been linked to inflammatory activity and immune regulation. While individual Trp metabolites have been associated with disease severity and treatment response, systems-level frameworks to define metabolic subtypes in IBD are lacking.

**Objective:** To identify reproducible Trp-related metabolic subtypes (“metabotypes”) in IBD and assess their association with disease activity, clinical outcomes, and early disease development.

**Design:** We applied unsupervised clustering to serum concentrations of 16 Trp-related metabolites in a discovery cohort of patients with IBD undergoing biologic induction therapy (n=134). Metabotypes were validated in three independent IBD cohorts (total n>2,800), a healthy reference population, and a prospective cohort of first-degree relatives at risk for Crohn’s disease. Associations with disease activity, longitudinal outcomes, and metabolic pathways were assessed using multivariable regression and survival analysis.

**Results:** Four reproducible metabotypes with distinct metabolite profiles were identified across cohorts: Low Kyna, High Kyna, High Quin, and Balanced. Low Kyna and High Quin metabotypes were consistently associated with increased inflammatory activity and adverse clinical outcomes, including increased risk of treatment escalation and disease progression.

Pathway-level analyses revealed alterations in NAD-related, lipid, and amino acid pathways between inflammatory metabotypes. A metabotype resembling inflammatory disease states was enriched in individuals who later developed Crohn’s disease in a prospective pre-disease cohort.

**Conclusion:** Trp-linked metabotypes define reproducible immunometabolic states in IBD that associate with disease activity and clinical outcomes and may precede disease onset. These findings provide a framework for metabolic stratification and biomarker-guided clinical trials targeting immunometabolic pathways.

**What is already known on this topic:** Tryptophan metabolism through the kynurenine pathway is a central immunometabolic axis in inflammatory bowel disease (IBD) and has been linked to inflammatory activity and immune regulation. Individual tryptophan metabolites have been associated with disease severity and treatment response, but their clinical utility for patient stratification remains limited. Systems-level approaches to define clinically meaningful metabolic subtypes in IBD are lacking.

**What this study adds:** We identify four reproducible tryptophan-related metabolic subtypes (“metabotypes”) that are consistently associated with disease activity across multiple independent IBD cohorts. Inflammation-associated metabotypes show distinct pathway-level alterations, including differences in NAD-related metabolism and broader metabolic programs. A metabotype resembling inflammatory disease states is detectable before clinical diagnosis in individuals who later develop Crohn’s disease.

**How this study might affect research, practice or policy:** Metabotype-based classification provides a framework for molecular stratification of patients in mechanistic studies and clinical trials targeting immunometabolic pathways. This approach may support biomarker-guided monitoring of disease activity and disease progression in IBD. Identification of preclinical metabolic states highlights the potential of metabolomics for early disease detection and prevention-oriented research strategies.

## Introduction

Systemic inflammation profoundly alters host metabolism and can be captured by serum metabolomic profiling in inflammatory bowel disease (IBD). Tryptophan degradation through the kynurenine pathway represents a central immunometabolic axis in IBD and is closely linked to inflammatory signalling and immune regulation.[1,2]. During inflammation, heightened IDO activity leads to depletion of Trp and accumulation of downstream kynurenine metabolites such as kynurenine (Kyn), kynurenic acid (Kyna), and quinolinic acid (Quin)[3]. A major metabolic aim of kynurenine pathway is to increase the de-novo NAD^+^ synthesis from tryptophan, restore cellular functions (e.g. mitochondrial respiration, DNA repair)[4], necessary to maintain the mucosal barrier[5].

Several downstream metabolites of the kynurenine pathway have been investigated individually understand the direct impact on intestinal immunity, including Kynurenic acid (KYNA)[6,7], xanthurenic acid[8] and quinolinic acid[9]. Quin is impaired due to transcriptional suppression of quinolinate phosphoribosyltransferase (QPRT), a rate-limiting enzyme in *de novo* nicotinamide adenine dinucleotide (NAD^+^) synthesis. The resulting bottleneck promotes Quin accumulation and contributes to local NAD^+^ depletion, redox imbalance, and immune dysregulation[10].

While individual Trp metabolites have been associated with disease severity or even therapy response in IBD, systems-level frameworks are needed to capture the heterogeneity and dynamism of metabolomic states in affected individuals [11–13]. The concept of metabotypes (metabolically defined subgroups with shared physiological or pathological features) has been proposed and applied to resolve functional variation within complex diseases [14].

Whether metabolomic stratification can reveal meaningful biological subtypes in IBD remains unclear. Here, we hypothesized that distinct, disease-relevant metabotypes exist within the Trp pathway that reflect underlying immunometabolic states. To test this, we applied unsupervised clustering to serum Trp metabolomic profiles from four well-characterized IBD cohorts, spanning different geographic regions, disease stages, and metabolomic platforms. We evaluated the association of resulting metabotypes with clinical disease activity, longitudinal outcomes, and pathway-level metabolic signatures across multiple cohorts, including a healthy reference population and a prospective pre-disease cohort, where an inflammatory metabotype was enriched prior to clinical diagnosis.

## Methods

### Cohort: Discovery cohort (BFU+)

The initial discovery cohort comprised a subset of 134 patients (52 CD and 82 UC) from the Biologics Follow-up Plus (BFU+) cohort, a prospective registry of patients with IBD recruited in Kiel, Germany. Serum samples were selected based on availability at the time of analysis and included paired samples at baseline (week 0), week 2, and week 14 following induction with advanced biological therapies. Clinical and demographic characteristics are summarized in mental Table 1. Additional details regarding cohort design have been previously described [15]. Therapy response was defined by total Mayo reduction of at least 50% with concurrent reduction in endoscopic Mayo (UC) and Crohn’s disease activity index (CDAI) reduction of at least 100 points with concurrent simple endoscopic score-CD (SES-CD) reduction of at least 50% (CD). The BFU+ study was approved by the local ethics committee in Kiel (D490/20, D489/20, A124/14, 156/03-2/13).

### Validation cohorts: IBSEN III, Suivitheque, MSCCR

Serum samples were obtained from adults enrolled in the Inflammatory Bowel Disease in Southern Norway III (IBSEN III) inception cohort [16]. Participants were sampled at baseline and categorized as CD, UC, IBD-unclassified (IBD-U), or symptomatic non-IBD. Metabolomic profiling was performed on 571 individuals (179 CD, 340 UC, 209 non-IBD, 26 IBD-U) for whom serum was available from biobanking at three of the participating clinical centers. The IBSEN III study was approved by the Southeastern regional ethics committee in Norway (ref # 2015/946 REK sør-øst B).

Data from the Suivitheque cohort were analyzed as part of this study. The cohort design and clinical framework have been previously reported by Michaudel et al. [17]. Patients with IBD undergoing routine clinical follow-up provided serum samples, which were analyzed in conjunction with harmonized metadata. Cases included 788 CD, 281 UC and 50 individuals without IBD.

A third validation cohort consisted of samples and data from the Mount Sinai Crohn’s and Colitis Registry (MSCCR), a prospective observational cohort study [18]. Study design and procedures of MSCCR have been previously described. The MSCCR was approved by the Institutional Review Board (IRB) of the Mount Sinai Hospital (HS# 11-01669). Samples were collected at baseline index endoscopy in this registry. For this study, samples from 1173 individuals included 470 CD, 374 UC and 329 non-IBD cases. Active endoscopic disease activity was defined by endoscopic Mayo of 0 or 1 (UC), or SES-CD of 4 or less (CD). Prospective longitudinal outcome data were extracted from structured electronic health record (HER) data through the Mount Sinai Data Warehouse. Details of the ascertainment of study outcomes have been previously described [19].

Pre-disease cohort: The CCC-GEM Project recruited asymptomatic FDRs of CD patients, aged between 6 and 35, and prospectively followed them to monitor for the onset of CD. At recruitment, participants were screened to exclude those with gastrointestinal symptoms or any diagnosis of IBD, celiac disease, or irritable bowel syndrome. All participants, and/or their guardians, provided written informed consent to participate in the study. The Mount Sinai Hospital Research Ethics Board (Toronto-Managing Center) and all local recruitment centers approved the study. Participants were contacted every six months to check for any potential new diagnoses of CD. If a participant reported a CD diagnosis, this was confirmed by the treating physician based on clinical, endoscopic, radiographic, and/or histological reports. A nested case-control cohort was identified to include individuals who developed CD (78 pre-CD subjects) along with up to four healthy matched control who remained disease-free during the observation period (311 healthy match control). Matching criteria included age, sex assigned at birth, geographic location, and time of recruitment.

### Healthy reference population: Diener et al. 2022

Data from a previously published healthy reference cohort (1569 individuals) described by Diener et al. were used as a comparator group [20]. Participants were enrolled in the Arivale Scientific Wellness Program and underwent deep phenotyping, including targeted serum metabolomics. Overall sample size was determined by availability of metabolomics data in participating cohorts rather than formal power calculations.

### Metabolomics analysis

Discovery: Samples were shipped frozen to Biocrates Life Sciences AG in Innsbruck, Austria, where they were kept frozen at −80°C until they were measured using their ‘Tryptophan Metabolism Assay’. Serum samples were analysed using targeted UHPLC–MS/MS with isotope-labelled internal standards according to validated protocols.

Serum samples from the IBSEN III cohort were analyzed using targeted LC-MS/MS with a panel for B vitamins and kynurenines (BEVITAL; <u>bevital.no</u>) as described in Midttun et al. [21].The targeted quantitative metabolomics workup for the Suivitheque cohort has also been previously described [17]. Samples from MSCCR were used for semi-quantitative untargeted metabolomics analysis performed by Metabolon, Inc. (Research Triangle Park, North Carolina, United States) [22,23]. Serum metabolomic measurements from the GEM project were performed with Metabolon using the Metabolon’s DiscoveryHD4™ Platform following manufacturer instructions as previously described (PMID: 40910526).

### Preprocessing and feature selection

The discovery cohort analysis focused on a targeted panel of tryptophan-related metabolites. These included: 3-OH-anthranilic acid (3OH-AA), 3-OH-kynurenine (3OH-Kyn), 5-OH-tryptophan (5OH-Trp), anthranilic acid (AA), kynurenic acid (Kyna), kynurenine (Kyn), picolinic acid (Pico), quinaldic acid (Quinald), quinolinic acid (Quin), xanthurenic acid (Xan), tryptophan (Trp), serotonin (Sero), indole-3-propionic acid (IPA), 5-hydroxyindoleacetic acid (5-HIAA), neopterin (Neopt), nicotinamide (NAM), and nicotinic acid.

Metabolites detected in fewer than 80% of samples were excluded. Nicotinic acid, while initially measured, was excluded due to excessive missingness (326 missing values, >20% of samples). Missing values were imputed using half of the lowest observed value. Data were log-transformed and standardized. Clustering analyses were performed on metabolites that passed initial preprocessing. All other cohorts used the same preprocessing workflow, with feature selection based on the discovery metabolite set.

### Unsupervised clustering and cluster selection

To identify metabolically distinct patient subgroups (metabotypes) in the discovery cohort, we evaluated multiple clustering algorithms and metrics. Multiple clustering algorithms were evaluated during exploratory analyses. The optimal number of clusters was determined using internal validation metrics assessing cluster stability and separation. Cluster stability was evaluated via resampling using the clusterboot procedure and multivariate assessment with the clValid package (0.7). Multiple clustering approaches were evaluated, and a four-cluster solution was selected based on stability metrics and biological interpretability. Considering both algorithmic consensus and previous reports linking low levels of Xan, Kyna and high levels of Quin to disease activity, we selected k = 4 as the final number of clusters. For the validation cohorts, we applied the final workflow as selected by the pipeline outlined above.

### Consensus clustering implementation

Final clustering was performed using consensus clustering to ensure stability and reproducibility. The input matrix consisted of log-transformed and standardized metabolite concentrations. Consensus clustering was performed using k-means with resampling. In each iteration, samples were subsampled while retaining all features. Consensus assignments at k = 4 were extracted and mapped back to the original dataset. Clusters were annotated based on their characteristic metabolite signatures as follows: Low Kyna, High Kyna, High Quin, and Balanced.

### Metabotype distribution and demographic associations

Normalized metabolite profiles were visualized using heatmaps grouped by metabotype and annotated by clinical covariates. Demographic and diagnostic features were compared across metabotypes using appropriate statistical tests. Disease activity indices were compared using linear mixed models adjusted for age and sex with correction for multiple testing. Mixed models included patient ID as a random intercept.

To identify traits enriched or depleted in specific metabotypes within the deeply phenotyped MSCCR cohort, we performed a structured enrichment analysis across a set of categorical clinical variables, including biologic-naïve status, family history, smoking status, current corticosteroid or probiotic use, Crohn’s disease behavior and location, ulcerative colitis superficial inflammation and extent, presence of other autoimmune diseases, disease duration, and patient-reported disease activity over the preceding six months as assessed by the Manitoba IBD index. For each variable, contingency tables were generated by metabotype. Global associations were tested using Fisher’s exact test with simulated p-values (10,000 permutations), and FDR correction was applied. Odds ratios (ORs) and approximate 95% confidence intervals were calculated based on observed and expected frequencies under the null hypothesis of independence. Significant results (FDR < 0.05) were displayed as a heatmap with OR magnitude and direction encoded by color. In IBSEN III, Fisher’s exact test with FDR adjustment was used to compare rates of aggressive disease between metabotypes.

### Longitudinal outcomes and survival analysis

Cox proportional hazards models were used to assess the association between metabotype and treatment escalation and disease-related outcomes. Metabotype was modelled as a categorical variable. The added predictive value of metabotype was assessed using likelihood ratio tests comparing models with and without metabotype. Endoscopic activity was incorporated as a covariate to distinguish metabotype associations that reflect independent predictive value from those driven primarily by concurrent disease activity. Global p-values were derived from likelihood ratio tests. Adjusted hazard ratios and cumulative incidence curves were estimated by metabotype.

### Pathway Enrichment Analysis

To assess pathway-level metabolic perturbations between the two high disease metabotypes (Low Kyna and High Quin), we performed gene set enrichment analysis (GSEA) using the fgsea package (1.30.0). For this pairwise comparison of metabotypes, we constructed linear models to estimate the association between metabolite abundance and metabotype assignment, adjusting for participant age and sex. Metabolites were ranked by model coefficient, and pathway-level enrichment was tested using curated sub-pathway annotations provided by Metabolon. Normalized enrichment scores (NES) and adjusted p-values (FDR) were used to identify pathways significantly perturbed between the metabotypes.

### Patient and public involvement

Patients and members of the public were not involved in the design, conduct, reporting, or dissemination plans of this research. The study was based on analysis of existing clinical cohorts and biobanked samples collected as part of routine clinical care and observational research programs. Findings from this work will inform future studies aimed at improving patient stratification and disease monitoring in inflammatory bowel disease.

## Results

### Metabotypes associate with disease activity across diverse clinical cohorts

Within the discovery cohort (BFU+), unsupervised clustering of 16 tryptophan-related metabolites revealed four stable patient subgroups differentially associated with disease activity, as measured by CRP, CDAI, and partial Mayo score (Fig. 1A, Fig. S1A). The metabotype exhibiting the highest disease activity was characterized by consistently decreased levels of the anti-inflammatory metabolites kynurenic acid (Kyna) and xanthurenic acid (Xan)[8]; this group was designated Low Kyna. In contrast, the High Kyna cluster showed elevated levels of Kyna and Xan and was associated with the lowest disease burden. A third group, High Quin, displayed moderately elevated disease activity and was defined by increased levels of quinolinic acid (Quin), kynurenine (Kyn), and neopterin (Neopt). The fourth cluster showed intermediate variation between metabolites and was therefore labeled Balanced.

**Figure 1.**
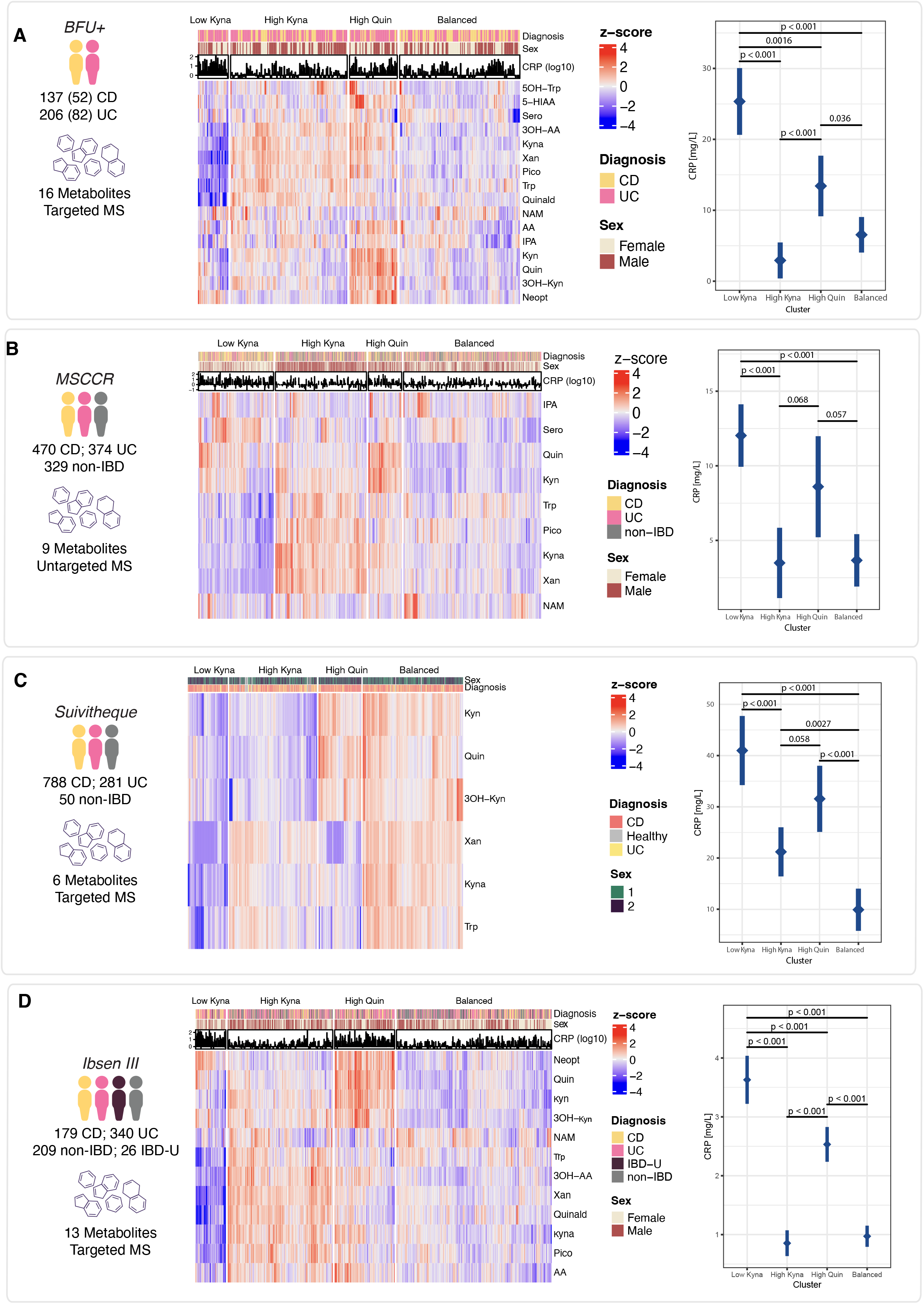
Metabotypes derived from tryptophan pathway metabolites are associated with disease activity across four IBD cohorts. **(A-D)** Consensus clustering (k = 4) of 16 serum tryptophan pathway metabolites identified four metabotypes in the discovery cohort and reproduced similar structures in Suivitheque, MSCCR, and IBSEN III. Clusters were named based on dominant metabolite signatures: Low Kyna, High Kyna, High Quin, and Balanced. Heatmaps display Z-score normalized metabolite intensities; top annotations indicate diagnosis, sex, and CRP levels.

Consensus clustering was independently applied to three additional IBD cohorts: Suivitheque, MSCCR, and IBSEN III (Fig. 1B-D, Fig. S1B-D). Despite variation in the available disease activity indices and metabolite measurement platforms, clustering with k=4 identified similar metabolic groupings and associations with disease activity. The Low Kyna group exhibited the highest clinical activity in all cohorts, while High Quin was associated with moderate inflammation, and the Balanced and High Kyna clusters generally showed lower clinical scores. Clustering was also performed on a cohort of healthy individuals, where no stable clusters related to tryptophan pathway metabolites were identified (Supplemental Figure 2), indicating that the observed metabotypes reflect disease-specific variation.

### Metabotype associations with clinical features and outcomes

Expanding beyond disease activity, we also found differences in distribution of sex and age of individuals across metabotypes. The Low Kyna group tended to contain more females, and the High Quin group was older compared to the other metabotypes (Supplemental Tables 1-4). Only the MSCCR cohort provided information on race, showing no evidence of differential enrichment between racial groups (Supplemental Table 3).

To delve deeper into the clinical features and longitudinal outcomes associated with each metabotype, we took advantage of the wealth of clinical data and sample size available from the MSCCR cohort. We report clinical variables exhibiting differential enrichment between the metabotypes (FDR < 0.05) (list of variables tested accompanied by statistical approach is listed in the methods section). Most importantly, we found that individuals classified as Low Kyna more likely received corticosteroids or probiotics at the time of sampling and were more likely to report “often” or “constantly” active disease over the previous 6-months in the Manitoba IBD index questionnaire [24]. Although corticosteroid use was significantly enriched in the Low Kyna cluster in the MSCCR cohort, the same cluster structure was observed in the IBSEN III inception cohort, where corticosteroid exposure was minimal at the time of sampling (Supplemental Table 5), suggesting that the enrichment reflects underlying disease activity rather than treatment effect. In addition, there was an enrichment for extensive colitis in High Quin-classed individuals with UC (Fig. 2A).

**Figure 2.**
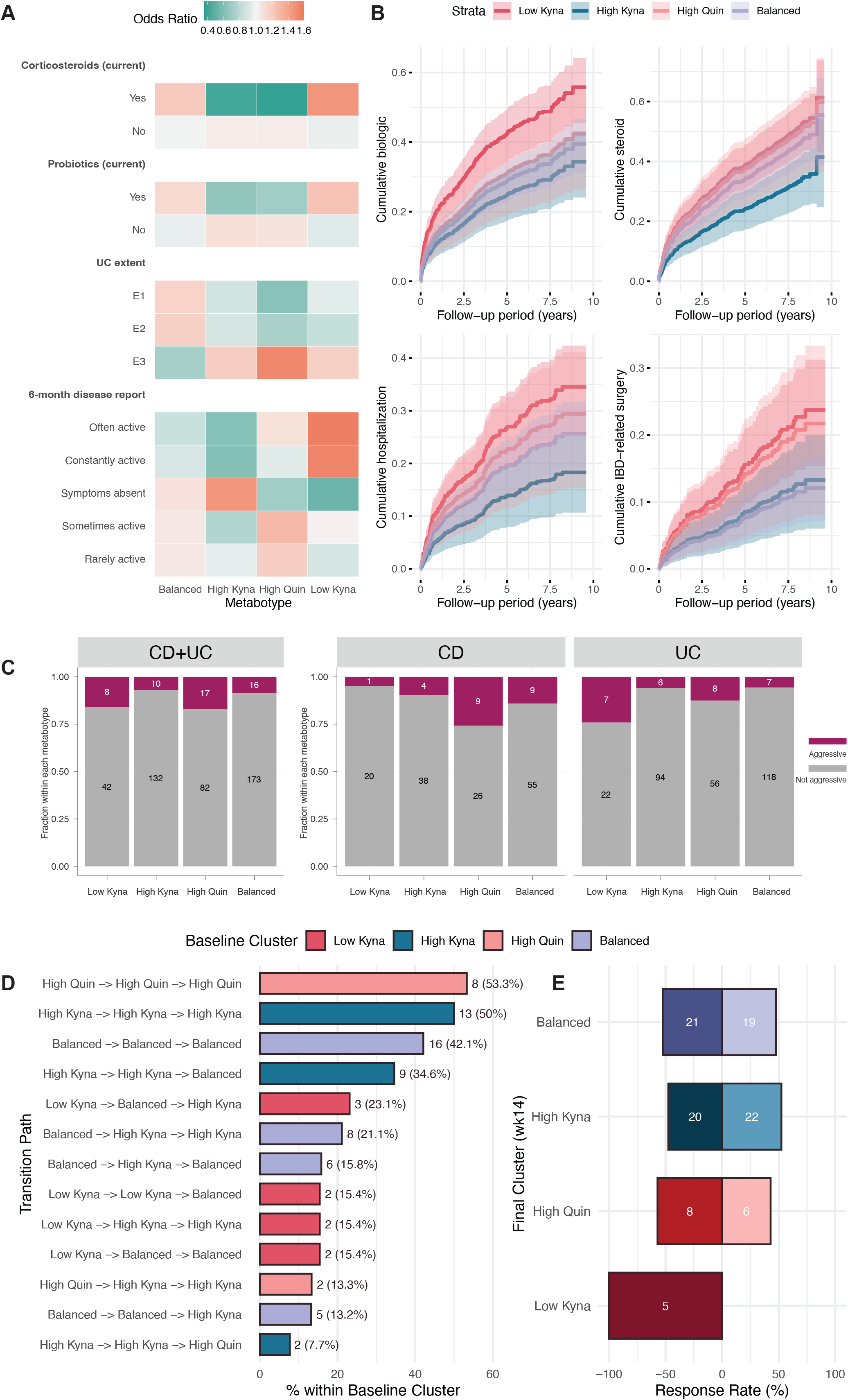
Metabolomic cluster assignment is associated with clinical features and longitudinal outcomes. **(A)** Odds ratios show enrichment of clinical features across metabotypes. **(B)** Survival curves for biologic initiation, novel corticosteroid use, IBD-related hospitalization, and IBD-related surgery by cluster (Cox models adjusted for age and sex). **(C)** High Quin showed the highest proportion of aggressive disease in the IBSEN III cohort. **(D)** Top 15 observed metabotype transitions from baseline, week 2 and week 14 for patients with metabolomics data at each of the three time points. **(E)** Final cluster assignment at week 14 and its relationship with therapy response. A,B: MSCCR; C: IBSEN III; D,E: Discovery cohort.

Next, we observed that cluster membership was associated with risk of initiating novel biologic or corticosteroid therapy, IBD-related hospitalization, IBD-related surgery, and a composite outcome of disease progression, defined as the occurrence of any of the aforementioned events (global p = 0.0018, 0.0062, 0.021, 0.020, and 0.0042, respectively; Fig. 2B and Supplemental Table 6). The largest effect sizes and strongest statistical evidence were found when comparing the hazards of the Low Kyna to the High Kyna metabotypes (Supplemental Table 6). These associations were attenuated after adjusting for baseline biologic treatment and endoscopic disease activity, except for the risk of novel corticosteroid use (p = 0.048; Supplemental Table 6). In contrast, in the independent inception cohort (IBSEN III), individuals in the High Quin group were more likely to experience a severe disease course the first year after diagnosis, consistent with a more severe clinical phenotype (Fig. 2C, Supplemental Table 7-8).

Lastly, the discovery cohort (BFU+) was used to assess within patient transitions between metabotypes. We first observed the transition path of metabotypes across three time points following biologic therapy initiation (baseline, week 2, week 14). Longitudinal sampling in the discovery cohort revealed frequent transitions between metabotypes over 14 weeks of treatment (Fig. 2D). Patients with lower risk metabotypes (High Kyna and Balanced) at baseline were most likely to remain in their metabotype throughout the period of observation, and transitions most often occurred between these two states. As the metabotype classification reflects disease activity, we analyzed the proportion of responders and non-responders at week 14 for each metabotype. All 5 individuals classified as Low Kyna at week 14 failed to achieve clinical response to therapy at week 14 (Fig. 2E).

### Pathway-level contrasts between inflammatory metabotypes

To investigate broader metabolic differences between the two metabotypes most enriched for disease activity (Low Kyna, High Quin), we performed pathway-level analysis in the MSCCR cohort, where the semi-targeted nature of the data allowed for a more complete picture of altered pathways beyond Trp metabolism. As de novo synthesis is a major metabolic outcome of Trp degradation along the Kyn pathway, we performed a focused analysis on NAD^+^-related metabolites. Among NAD metabolites quantified in the MSCCR cohort, 1-methyl-NAM was lowest in Low Kyna, while 2-PY was highest in High Quin (Figure 3, Supplemental Table 9); trigonelline and NAM did not differ. Expanding beyond the Trp-related metabolites, pathway enrichment using the entire set of metabolites measured revealed broad changes in amino acid and lipid metabolism between the High Quin and Low Kyna groups (Fig. 3B).

**Figure 3.**
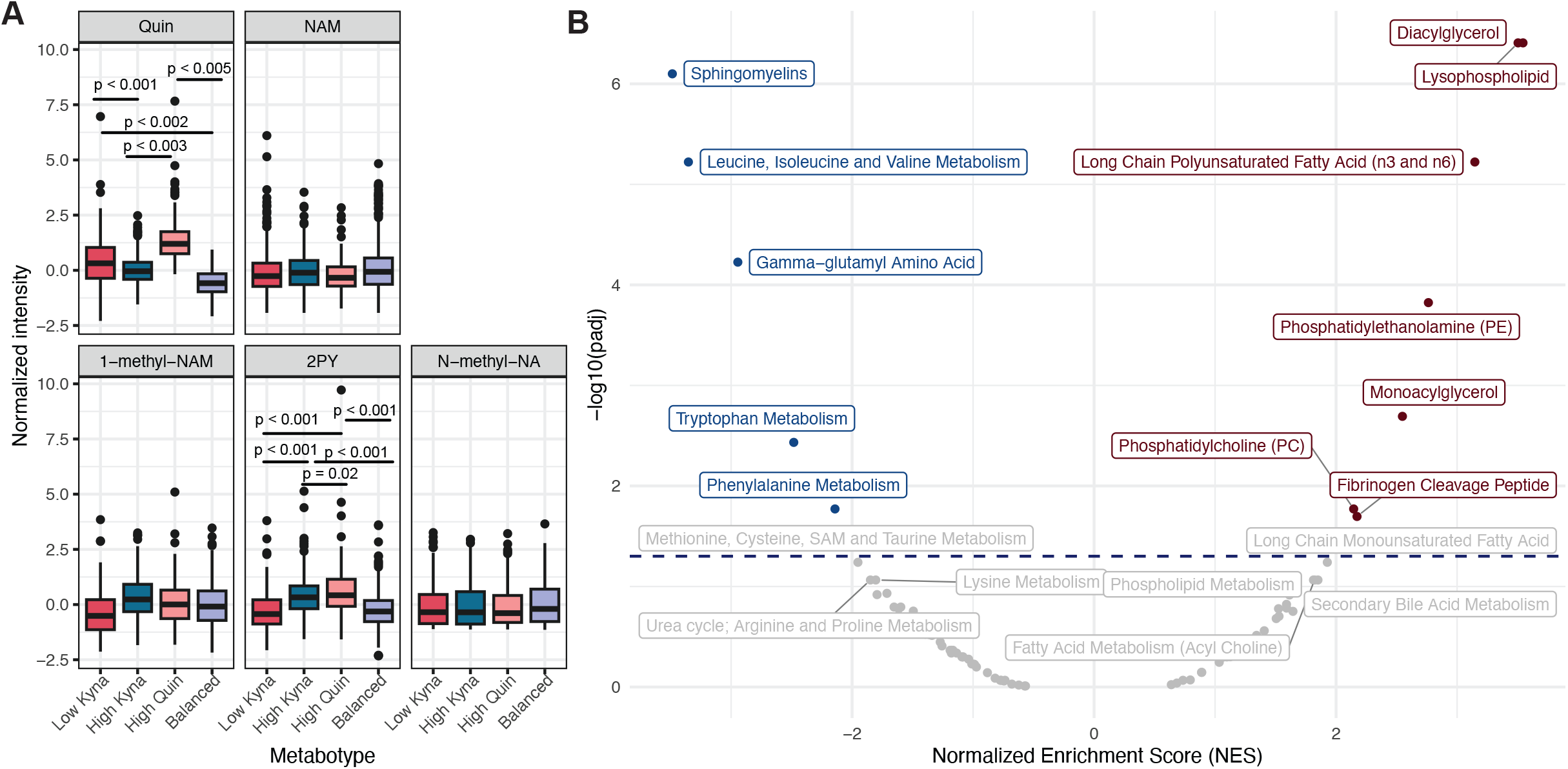
Pathway-level and metabolite-level differences between High Quin and Low Kyna metabotypes in the MSCCR cohort. **(A)** Serum concentrations of NAD^+^-related metabolites across metabotypes. Although it was one of the features used for clustering, Quin is included here for reference. **(B)** Pathway-level enrichment analysis (High Quin vs Low Kyna) based on normalized enrichment scores (NES) and adjusted p-values. Positive NES values indicate pathways enriched in Low Kyna relative to High Quin, while negative NES values indicate pathways enriched in High Quin.

### Distinct metabolic subtype enrichment in future CD cases

Lastly, to assess whether disease-associated metabolic states precede clinical onset, we applied consensus clustering to serum metabolites from a prospective cohort of first-degree relatives of patients with CD in the GEM cohort (Fig. 4)[25,26]. This cohort included individuals who remained healthy and those who progressed to CD (“pre-CD”) during follow-up. Clustering identified four groups with distinct metabolic profiles compared to those observed in established IBD cohorts. One cluster (Cluster 1) exhibited a composite signature resembling features of both the Low Kyna and High Quin metabotypes. Cluster 1 was enriched for inflammatory markers, including CRP, fecal calprotectin (FCP), and lactulose-to-mannitol ratio (LMR, representing intestinal permeability) (Figure 4B-D), and contained higher proportions of individuals who later developed CD (Figure 4E). Time to diagnosis was shorter in Cluster 1 than Cluster 2, but the difference did not reach statistical significance (p = 0.075). However, this inflammatory phenotype was absent in a healthy reference cohort (Supplemental Figure 1), underscoring its disease relevance.

**Figure 4.**
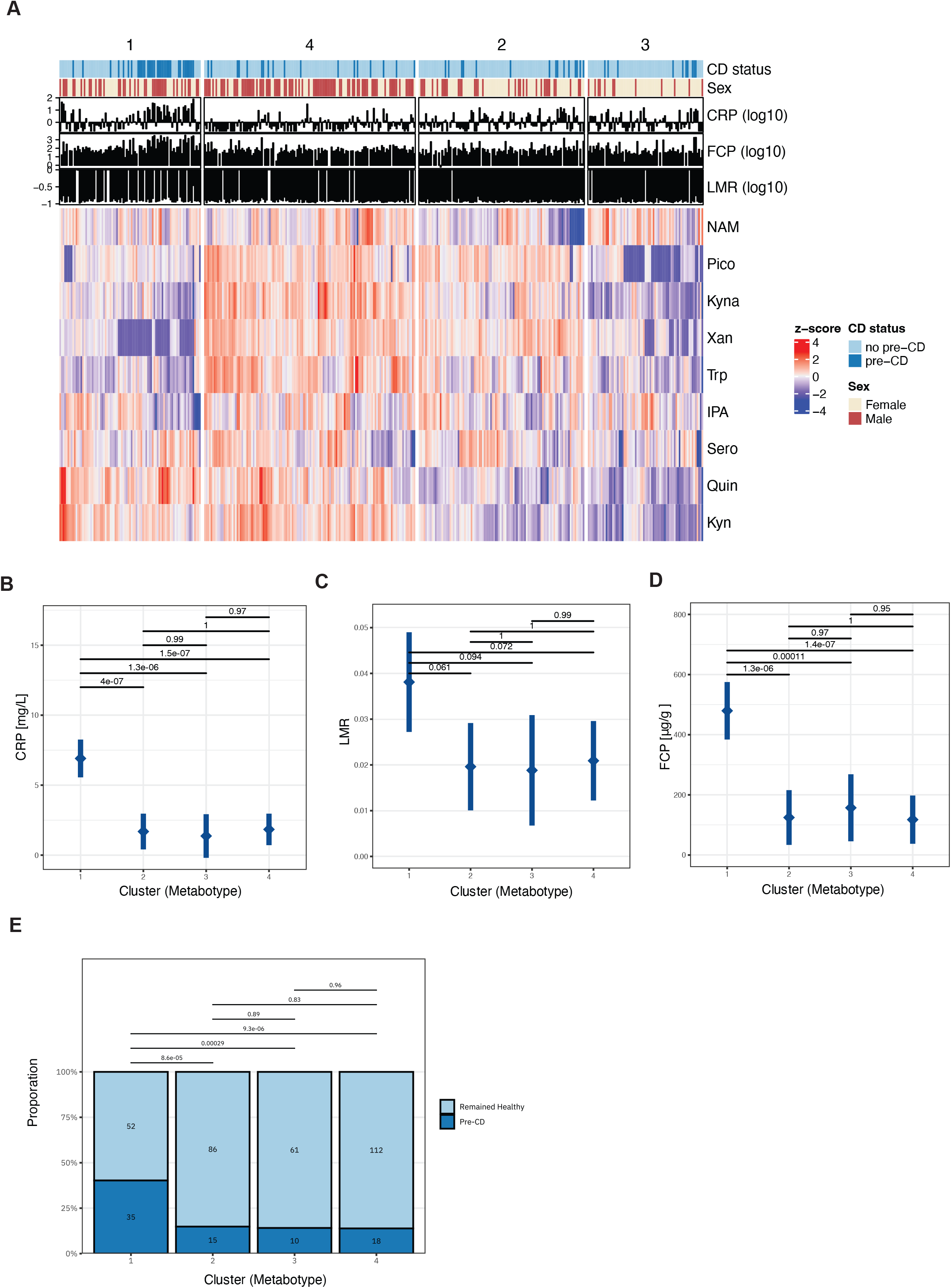
Distinct metabolic subtype enrichment in future CD cases in the GEM cohort. (A) Heatmap of serum metabolite z-scores across four consensus clusters. Cluster annotation bars indicate CD status (pre-CD vs remained healthy) and sex. (B-D) Inflammatory markers include CRP (B), lactulose-to-mannitol ratio (LMR, C), and fecal calprotectin (FCP, D) across clusters, with error bars representing mean ± standard error. (E) Proportion of individuals who remained healthy versus progressed to CD (“pre-CD”) in each cluster.

## Discussion

Trp catabolism, primarily along the kynurenine pathway, emerged as a key immunometabolic axis in IBD, yet few studies have evaluated its potential to define biologically or clinically meaningful patient subgroups [27]. We identified four reproducible tryptophan-related metabotypes associated with disease activity across independent IBD cohorts. The inflammatory metabotypes showed distinct biochemical and metabolic profiles. These findings suggest that distinct metabolic programs may be associated with increased disease severity, with potential implications for disease pathogenesis and treatment response. Although metabotypes were associated with adverse clinical outcomes, the effects were not independent of quantitative measure of disease activity (e.g. endoscopic disease activity). However, they offer orthogonal insight into the biochemical consequences of inflammation, suggesting distinct immunometabolic programs that are reproducible across disease stages, cohorts, and platforms and could be feasible for non-invasive activity assessment or longitudinal monitoring. This cross-cohort reproducibility highlights the robustness of the metabotype classification despite differences in geography, clinical scoring systems, sample processing, and assay platforms. The presence of the metabotypes in the IBSEN III inception cohort suggests that these phenotypes are established early in the disease course, remain stable across disease history, and are unlikely to reflect treatment artifacts.

To better understand the biological distinctions between the two inflammation-associated metabotypes High Quin and Low Kyna, we examined differences in pathway-level metabolic activity. Given the established link between tryptophan catabolism and NAD^+^ biosynthesis, we assessed whether markers of the NAD^+^ pathway differed between metabotypes. Importantly, these metabotypes reflect two pathophysiological principles that we have previously described. Reduced kynurenic acid (Kyna) and xanthurenic acid (Xana) have been associated with increased disease activity and risk of flare in IBD, and modulation of the key enzyme involved in their synthesis (aminoadipate aminotransferase, ADAT) has been shown to ameliorate disease activity in experimental colitis [8] and was also linked to disease activity in rheumatoid arthritis[28]. In addition to reduced Kyna, we observed increased quinolinic acid (Quin) as a metabotype associated with disease activity. Quin serves as a precursor for NAD^+^ via the de novo synthesis pathway, and its accumulation has been linked to impaired NAD^+^ production during intestinal inflammation through suppression of QPRT. Consistent with this framework, our targeted metabolomics panel captured key NAD-related metabolites, supporting the interpretation that alterations in NAD metabolism accompany inflammatory metabotypes.

[8][28][15][29]

Although we hypothesized that increased Quin levels in the serum of patients assigned to the High Quin metabotype might reflect impaired *de novo* NAD^+^ synthesis, our metabolite panel did not provide direct evidence of reduced NAD^+^ biosynthetic capacity. However, the serum elevation of 2-PY may suggest enhanced downstream NAD^+^ catabolism, potentially driven by inflammatory NAD^+^ consumption [30]. [31,32]These findings are consistent with previous observations of reduced fecal NAM levels[33] and reduced epithelial NAD^+^ levels in the intestinal mucosa of IBD patients[5].

Broader pathway analysis comparing High Quin and Low Kyna revealed coordinated shifts across multiple metabolic processes. In Low Kyna metabotypes, amino acid metabolism and steroid hormone biosynthesis pathways were among the most significantly downregulated, while choline and benzoate pathways were enriched. In High Quin, membrane lipid classes were enriched, alongside elevated 2-PY, a marker of NAD^+^ clearance. These findings support distinct metabolic responses associated with inflammatory disease states.

Finally, we observed a distinct metabolic cluster enriched in individuals who progressed to CD in the GEM prospective preclinical cohort. This cluster shared features with both Low Kyna and High Quin and was enriched for inflammatory markers including CRP and FCP, suggesting that the underlying pathophysiologic processes may be active before clinical disease onset. Although this finding does not currently support prospective risk prediction, it indicates that early metabolic dysregulation may mark a primed immune state. The absence of a similar cluster structure in healthy individuals further supports the disease specificity of these metabolic programs.

Our study has several limitations. Although reproducibility across cohorts mitigated potential effects of sample handling and platform differences, we lacked dietary, immune, and transcriptomic data to define the upstream drivers of each metabotype. These metabolic patterns may therefore reflect immune-metabolic states not fully captured by conventional inflammatory markers. Potential sources of bias include differences in metabolomics platforms across cohorts and residual confounding from unmeasured variables.

Our study provides a molecular framework linking tryptophan metabolism to clinically relevant disease states in inflammatory bowel disease. Emerging clinical trials targeting amino acid metabolism and NAD-related pathways highlight the translational potential of modulating immunometabolic programs in chronic inflammation [34–36]. In this context, our data-driven metabotype framework may support biomarker-guided patient stratification and facilitate patient selection in future interventional studies targeting immunometabolic pathways.

## Data Availability

All data produced in the present study are available upon reasonable request to the authors
All data produced in the present work are contained in the manuscript
All data produced are available online at

https://www.ikmb.uni-kiel.de

## Acknowledgment

This work was supported in part through the Minerva computational and data resources and staff expertise provided by Scientific Computing and Data at the Icahn School of Medicine at Mount Sinai. This work is supported in part through the use of the research platform AI-Ready Mount Sinai (AIR·MS) and the expertise provided by the team at the Hasso Plattner Institute for Digital Health at Mount Sinai (HPI·MS). We gratefully acknowledge the excellent work of our technical assistants Christina Nimke, Janina Ohrndorf, Meike Hansen, Ronja Möhring, Sophie Reiher, Sabine Kock, Tanja Klostermeier, Stefanie Rentzow and Dorina Ölsner. We would further like to extend our sincere gratitude to the patients who participated in this study, without whom our research would not be possible.

ChatGPT (OpenAI) was used as a language support tool for editing and formatting text during manuscript preparation. The authors retained full responsibility for all scientific content, analyses, and conclusions, and all AI-generated suggestions were reviewed and approved by the authors.

## References

1 Xue C, Li G, Zheng Q, et al. Tryptophan metabolism in health and disease. Cell Metab. 2023;35:1304–26. doi: 10.1016/j.cmet.2023.06.004

2 Hashimoto T, Perlot T, Rehman A, et al. ACE2 links amino acid malnutrition to microbial ecology and intestinal inflammation. Nature. 2012;487:477–81. doi: 10.1038/nature11228

3 Agus A, Planchais J, Sokol H. Gut Microbiota Regulation of Tryptophan Metabolism in Health and Disease. Cell Host Microbe. 2018;23:716–24.

4 Katsyuba E, Mottis A, Zietak M, et al. De novo NAD^+^ synthesis enhances mitochondrial function and improves health. Nature. 2018;563:354–9. doi: 10.1038/s41586-018-0645-6

5 Novak EA, Crawford EC, Mentrup HL, et al. Epithelial NAD^+^ depletion drives mitochondrial dysfunction and contributes to intestinal inflammation. Front Immunol. 2023;14. doi: 10.3389/fimmu.2023.1231700

6 Wang D, Wang W, Bing X, et al. GPR35-mediated kynurenic acid sensing contributes to maintenance of gut microbiota homeostasis in ulcerative colitis. FEBS Open Bio. 2023;13:1415–33. doi: 10.1002/2211-5463.13673

7 Wang J, Simonavicius N, Wu X, et al. Kynurenic Acid as a Ligand for Orphan G Protein-coupled Receptor GPR35 *. Journal of Biological Chemistry. 2006;281:22021–8. doi: 10.1074/jbc.M603503200

8 Michaudel C, Danne C, Agus A, et al. Rewiring the altered tryptophan metabolism as a novel therapeutic strategy in inflammatory bowel diseases. Gut. 2023;72:1296–307. doi: 10.1136/gutjnl-2022-327337

9 McReynolds M, Alsaadi A, Welz L, et al. Tracing NAD^+^ metabolism uncovers adaptive coordination between host and microbiome during colitis. 2025.

10 Minhas PS, Liu L, Moon PK, et al. Macrophage de novo NAD^+^ synthesis specifies immune function in aging and inflammation. Nat Immunol. 2019;20:50–63. doi: 10.1038/s41590-018-0255-3

11 Nikolaus S, Schulte B, Al-Massad N, et al. Increased Tryptophan Metabolism Is Associated With Activity of Inflammatory Bowel Diseases. Gastroenterology. 2017;153:1504–1516.e2. doi: 10.1053/j.gastro.2017.08.028

12 Harris DMM, Szymczak S, Schuchardt S, et al. Tryptophan degradation as a systems phenomenon in inflammation-an analysis across 13 chronic inflammatory diseases. EBioMedicine. 2024;102:105056. doi: 10.1016/j.ebiom.2024.105056

13 Sigall Boneh R, van der Kruk N, Wine E, et al. Tryptophan metabolites profile predict remission with dietary therapy in pediatric Crohn’s disease. Therap Adv Gastroenterol. 2025;18. doi: 10.1177/17562848251323004

14 Semmar N. Metabotype Concept: Flexibility, Usefulness and Meaning in Different Biological Populations. Metabolomics. InTech 2012.

15 Welz L, Harris DM, Kim N-M, et al. A metabolic constraint in the kynurenine pathway drives mucosal inflammation in IBD. medRxiv. Published Online First: 8 August 2024. doi: 10.1101/2024.08.08.24311598

16 Kristensen VA, Opheim R, Perminow G, et al. Inflammatory bowel disease in South-Eastern Norway III (IBSEN III): a new population-based inception cohort study from South-Eastern Norway. Scand J Gastroenterol. 2021;56:899–905. doi: 10.1080/00365521.2021.1922746

17 Michaudel C, Danne C, Agus A, et al. Rewiring the altered tryptophan metabolism as a novel therapeutic strategy in inflammatory bowel diseases. Gut. 2023;72:1296–307. doi: 10.1136/gutjnl-2022-327337

18 Kosoy R, Kim-Schulze S, Rahman A, et al. Deep Analysis of the Peripheral Immune System in IBD Reveals New Insight in Disease Subtyping and Response to Monotherapy or Combination Therapy. Cell Mol Gastroenterol Hepatol. 2021;12:599–632. doi: 10.1016/j.jcmgh.2021.03.012

19 Bourgonje AR, Ibing S, Livanos AE, et al. Distinct perturbances in metabolic pathways associate with disease progression in inflammatory bowel disease. J Crohns Colitis. 2025;19. doi: 10.1093/ecco-jcc/jjaf082

20 Brandão-Lima PN, de Carvalho GB, Payolla TB, et al. Circulating microRNA Related to Cardiometabolic Risk Factors for Metabolic Syndrome: A Systematic Review. Metabolites. 2022;12:1044. doi: 10.3390/metabo12111044

21 Midttun È, Hustad S, Ueland PM. Quantitative profiling of biomarkers related to B-vitamin status, tryptophan metabolism and inflammation in human plasma by liquid chromatography/tandem mass spectrometry. Rapid Communications in Mass Spectrometry. 2009;23:1371–9. doi: 10.1002/rcm.4013

22 Ford L, Kennedy AD, Goodman KD, et al. Precision of a Clinical Metabolomics Profiling Platform for Use in the Identification of Inborn Errors of Metabolism. J Appl Lab Med. 2020;5:342–56. doi: 10.1093/jalm/jfz026

23 Bridgewater BR EA. High Resolution Mass Spectrometry Improves Data Quantity and Quality as Compared to Unit Mass Resolution Mass Spectrometry in High-Throughput Profiling Metabolomics. Journal of Postgenomics Drug & Biomarker Development. 2014;04. doi: 10.4172/2153-0769.1000132

24 Clara I, Lix LM, Walker JR, et al. The Manitoba IBD Index: Evidence for a New and Simple Indicator of IBD Activity. Am J Gastroenterol. 2009;104:1754–63. doi: 10.1038/ajg.2009.197

25 Turner D, Kenigsberg S, Focht G, et al. Preclinical stages of Crohn’s disease defined by faecal calprotectin in asymptomatic first-degree relatives: screening framework for prevention trials. Gut. 2025;gutjnl-2025-336368. doi: 10.1136/gutjnl-2025-336368

26 Xue M, Lee S-H, Shao J, et al. Metabolomics reveal distinct molecular pathways associated with future risk of Crohn’s Disease. Gut Microbes. 2025;17:2546998. doi: 10.1080/19490976.2025.2546998

27 Zheng X, Zhu Y, Zhao Z, et al. The role of amino acid metabolism in inflammatory bowel disease and other inflammatory diseases. Front Immunol. 2023;14:1284133. doi: 10.3389/fimmu.2023.1284133

28 Moulin D, Millard M, Taïeb M, et al. Counteracting tryptophan metabolism alterations as a new therapeutic strategy for rheumatoid arthritis. Ann Rheum Dis. 2023;83:312–23. doi: 10.1136/ard-2023-224014

29 Park J, Shin EJ, Kim TH, et al. Exploring NNMT: from metabolic pathways to therapeutic targets. Arch Pharm Res. 2024;47:893–913. doi: 10.1007/s12272-024-01519-9

30 Gerner RR, Klepsch V, Macheiner S, et al. NAD metabolism fuels human and mouse intestinal inflammation. Gut. 2018;67:1813–23. doi: 10.1136/gutjnl-2017-314241

31 Chen C, Tao M, Wang L, et al. Nicotinamide N-methyltransferase mediates redox regulation and colonic epithelial barrier impairment by SIRT1/PPAR-γ/NLRP6/NRF2 axis in inflammatory bowel disease. Int Immunopharmacol. 2026;168:115882. doi: 10.1016/j.intimp.2025.115882

32 Hou L, Du J, Li J, et al. 1-methylnicotinamide attenuated inflammation and regulated flora in Necrotizing enterocolitis. PLoS One. 2025;20. doi: 10.1371/journal.pone.0324068

33 Aoyama K, Yamamura R, Katsurada T, et al. Decreased Fecal Nicotinamide and Increased Bacterial Nicotinamidase Gene Expression in Ulcerative Colitis Patients. Inflamm Bowel Dis. 2025;31:2815–25. doi: 10.1093/ibd/izaf092

34 Christen S, Redeuil K, Goulet L, et al. The differential impact of three different NAD^+^ boosters on circulatory NAD and microbial metabolism in humans. Nat Metab. 2026;8:62–73. doi: 10.1038/s42255-025-01421-8

35 Norheim KL, Ben Ezra M, Heckenbach I, et al. Effect of nicotinamide riboside on airway inflammation in COPD: a randomized, placebo-controlled trial. Nat. Aging. 2024.

36 Schreiber S, Waetzig GH, López-Agudelo VA, et al. Nicotinamide modulates gut microbial metabolic potential and accelerates recovery in mild-to-moderate COVID-19. Nat Metab. Published Online First: 12 May 2025. doi: 10.1038/s42255-025-01290-1

