## Supplemental Files for "Tryptophan pathway metabotypes associate with disease activity and immune-metabolic dysfunction in inflammatory bowel disease"

**Supplemental filesx**

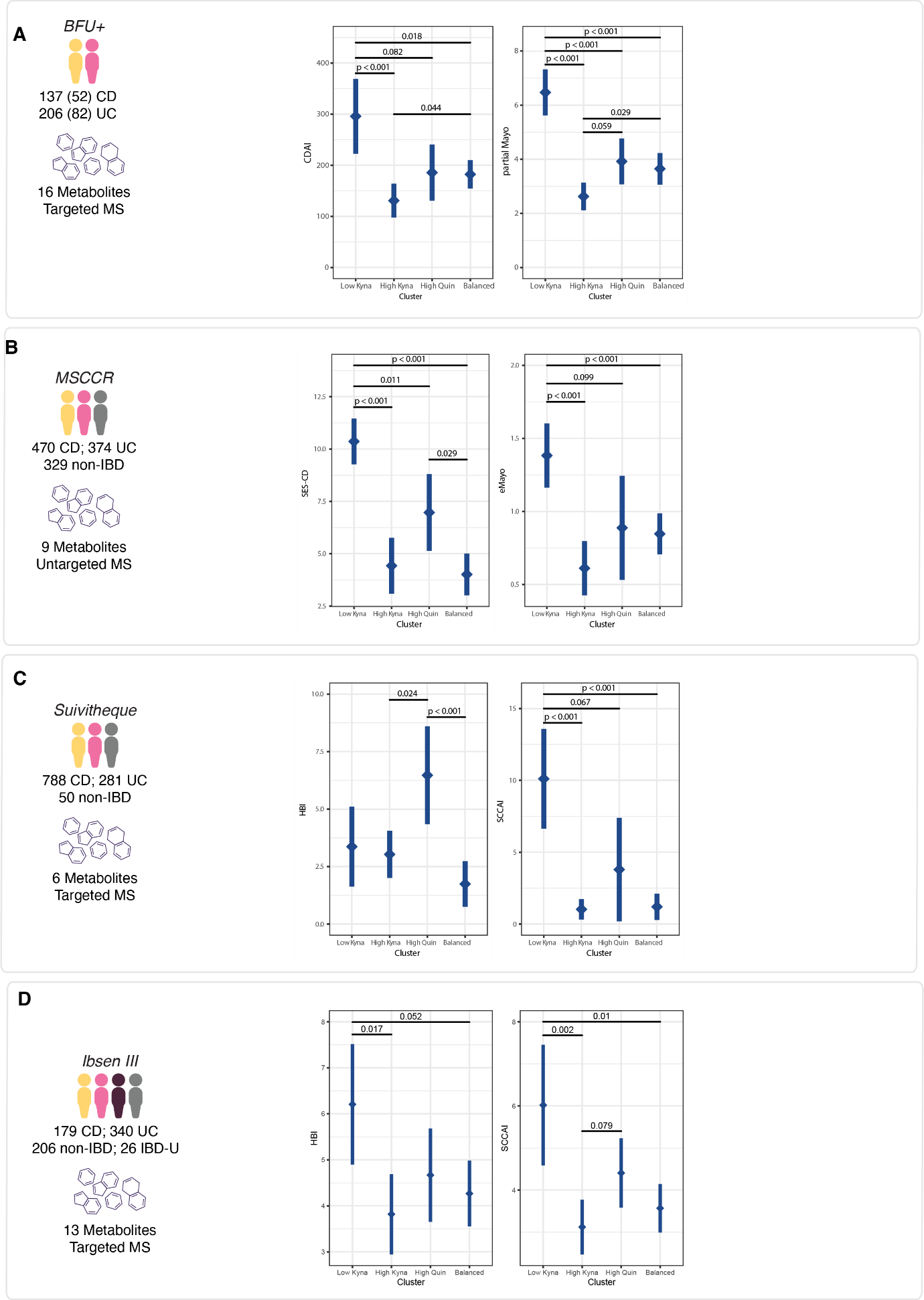

**Supplemental Figure 1.** Clinical and endoscopic disease scores for each individual cohort and their respective association with tryptophan metabotypes.

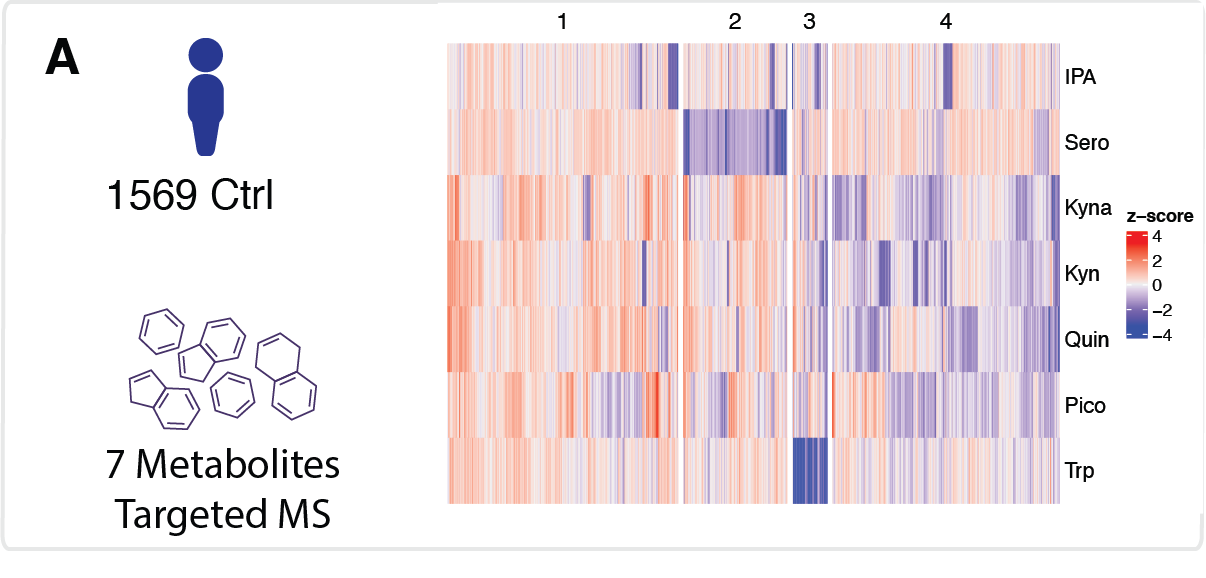

**Supplemental Figure 2.** No clear corollaries to the metabotypes observed in the IBD cohorts were recovered in the healthy reference cohort.

**Supplemental Table 1.** Demographics overview for BFU+. Age is provided in years.

|  | **level** | **Low Kyna** | **High Kyna** | **High Quin** | **Balanced** | **p** |
| --- | --- | --- | --- | --- | --- | --- |
| **n** |  | 21 | 37 | 21 | 55 |  |
| **Age (median [IQR])** |  | 30.00 [25.00, 36.00] | 36.00 [27.00, 48.00] | 58.00 [27.00, 72.00] | 32.00 [25.00, 45.50] | 0.008 |
| **Sex (%)** | Female | 13 (61.9) | 15 (40.5) | 8 (38.1) | 27 (49.1) | 0.354 |
|  | Male | 8 (38.1) | 22 (59.5) | 13 (61.9) | 28 (50.9) |  |
| **Diagnosis (%)** | CD | 5 (23.8) | 10 (27.0) | 7 (33.3) | 30 (54.5) | 0.017 |
|  | UC | 16 (76.2) | 27 (73.0) | 14 (66.7) | 25 (45.5) |  |

**Supplemental Table 2.** Demographics overview for Suivitheque. Age is provided in years.

|  | **level** | **Low Kyna** | **High Kyna** | **High Quin** | **Balanced** | **p** |
| --- | --- | --- | --- | --- | --- | --- |
| **n** |  | 165 | 363 | 177 | 414 |  |
| **Age (median [IQR])** |  | 39.53 [32.70, 50.03] | 38.28 [31.78, 49.74] | 41.11 [32.53, 56.20] | 40.90 [32.09, 55.05] | 0.051 |
| **Sex (%)** | Female | 105 (63.6) | 158 (48.3) | 108 (61.0) | 181 (45.2) | <0.001 |
|  | Male | 60 (36.4) | 169 (51.7) | 69 (39.0) | 219 (54.8) |  |
| **Diagnosis (%)** | CD | 126 (76.4) | 216 (59.5) | 146 (82.5) | 300 (72.5) | <0.001 |
|  | UC | 39 (23.6) | 111 (30.6) | 31 (17.5) | 100 (24.2) |  |
|  | non-IBD | 0 (0.0) | 36 (9.9) | 0 (0.0) | 14 (3.4) |  |

**Supplemental Table 3.** Demographics overview for MSCCR. Age is provided in years.

|  | **level** | **Low Kyna** | **High Kyna** | **High Quin** | **Balanced** | **p** |
| --- | --- | --- | --- | --- | --- | --- |
| **n** |  | 261 | 317 | 116 | 479 |  |
| **Age (median [IQR])** |  | 40.00 [31.00, 56.00] | 51.00 [37.00, 59.00] | 52.50 [36.75, 62.00] | 48.00 [33.00, 56.00] | <0.001 |
| **Sex (%)** | Female | 183 (70.1) | 65 (20.5) | 44 (37.9) | 270 (56.4) | <0.001 |
|  | Male | 78 (29.9) | 252 (79.5) | 72 (62.1) | 209 (43.6) |  |
| **Diagnosis (%)** | CD | 146 (55.9) | 103 (32.5) | 51 (44.0) | 170 (35.5) | <0.001 |
|  | UC | 72 (27.6) | 107 (33.8) | 26 (22.4) | 169 (35.3) |  |
|  | non-IBD | 43 (16.5) | 107 (33.8) | 39 (33.6) | 140 (29.2) |  |
| **Race (%)** | AI/AN | 2 (0.8) | 2 (0.6) | 0 (0.0) | 1 (0.2) | 0.492 |
|  | Asian | 5 (1.9) | 9 (2.8) | 3 (2.6) | 12 (2.5) |  |
|  | Black | 31 (11.9) | 25 (7.9) | 7 (6.0) | 37 (7.7) |  |
|  | Multiracial | 8 (3.1) | 6 (1.9) | 1 (0.9) | 9 (1.9) |  |
|  | Other | 22 (8.4) | 20 (6.3) | 10 (8.6) | 32 (6.7) |  |
|  | Refused | 0 (0.0) | 2 (0.6) | 1 (0.9) | 0 (0.0) |  |
|  | White | 193 (73.9) | 253 (79.8) | 94 (81.0) | 388 (81.0) |  |

**Supplemental Table 4.** Demographics overview for IBSEN III.

|  | **level** | **Low Kyna** | **High Kyna** | **High Quin** | **Balanced** | **p** |
| --- | --- | --- | --- | --- | --- | --- |
| **n** |  | 65 | 226 | 129 | 334 |  |
| **Age (median [IQR])** |  | 34 (27, 47) | 31 (25, 43) | 40 (30, 56) | 32 (26, 44) | <0.001 |
| **Sex (%)** | Female | 42 (65%) | 93 (41%) | 48 (37%) | 187 (56%) | <0.001 |
|  | Male | 23 (35%) | 133 (59%) | 81 (63%) | 147 (44%) |  |
| **Diagnosis (%)** | CD | 25 (38%) | 47 (21%) | 36 (28%) | 71 (21%) | <0.001 |
|  | UC | 33 (51%) | 104 (46%) | 70 (54%) | 133 (40%) |  |
|  | IBD-U | 2 (3.1%) | 6 (2.7%) | 5 (3.9%) | 13 (3.9%) |  |
|  | Non-IBD | 5 (7.7%) | 69 (31%) | 18 (14%) | 117 (35%) |  |

**Supplemental Table 5.** Overview of the number of patients within the IBSEN III cohort who were taking a therapy at the time of sampling.

| **Drug** | **CD, N = 179** | **Ctrl, N = 209** | **IBD-U, N = 26** | **UC, N = 340** |
| --- | --- | --- | --- | --- |
| Acetylsalicylic acid | 4 (2.2%) | 3 (1.4%) | 1 (3.8%) | 9 (2.6%) |
| Statins | 9 (5.0%) | 5 (2.4%) | 1 (3.8%) | 17 (5.0%) |
| Salazopyrin | 0 (0%) | 1 (0.5%) | 0 (0%) | 0 (0%) |
| 5-ASA | 6 (3.4%) | 0 (0%) | 1 (3.8%) | 61 (18%) |
| Prednisolon | 4 (2.2%) | 2 (1.0%) | 0 (0%) | 14 (4.1%) |
| Local treatment w steroids | 2 (1.1%) | 0 (0%) | 0 (0%) | 2 (0.6%) |
| TNF inhibitor | 6 (3.4%) | 0 (0%) | 2 (7.7%) | 4 (1.2%) |
| Imurel | 5 (2.8%) | 1 (0.5%) | 0 (0%) | 2 (0.6%) |
| Methotrexat | 1 (0.6%) | 1 (0.5%) | 0 (0%) | 2 (0.6%) |
| Other drugs | 65 (36%) | 59 (28%) | 8 (31%) | 99 (29%) |

**Supplemental Table 6.** Odds ratios and confidence intervals for the assessment of adverse outcomes in MSCCR. Two model results are presented: with (full) and without (base) adjustment for endoscopic disease activity and biologic status at the time of sampling. The outcomes tracked were introduction of new biologic (biol) or steroid, IBD-related hospitalization (hosp), IBD-related surgical procedure and finally hazards ratios for any of the aforementioned events (any).

| **Outcome** | **Contrast** | **HR** | **CI** | **p-val** | **model** |
| --- | --- | --- | --- | --- | --- |
| **biol** | Low Kyna / High Kyna | 1.94 | (1.21–3.12) | 0.00184 | base |
| **biol** | Low Kyna / High Quin | 1.48 | (0.8–2.73) | 0.348 | base |
| **biol** | Low Kyna / Balanced | 1.63 | (1.11–2.39) | 0.00622 | base |
| **biol** | High Kyna / High Quin | 0.76 | (0.41–1.42) | 0.681 | base |
| **biol** | High Kyna / Balanced | 0.84 | (0.54–1.29) | 0.726 | base |
| **biol** | High Quin / Balanced | 1.1 | (0.61–1.98) | 0.976 | base |
| **steroid** | Low Kyna / High Kyna | 1.77 | (1.12–2.82) | 0.00798 | base |
| **steroid** | Low Kyna / High Quin | 1.04 | (0.6–1.8) | 0.997 | base |
| **steroid** | Low Kyna / Balanced | 1.17 | (0.81–1.7) | 0.696 | base |
| **steroid** | High Kyna / High Quin | 0.59 | (0.34–1.02) | 0.0637 | base |
| **steroid** | High Kyna / Balanced | 0.66 | (0.44–0.99) | 0.0456 | base |
| **steroid** | High Quin / Balanced | 1.12 | (0.68–1.87) | 0.935 | base |
| **hosp** | Low Kyna / High Kyna | 2.1 | (1.11–3.96) | 0.0153 | base |
| **hosp** | Low Kyna / High Quin | 1.22 | (0.58–2.56) | 0.906 | base |
| **hosp** | Low Kyna / Balanced | 1.43 | (0.87–2.35) | 0.243 | base |
| **hosp** | High Kyna / High Quin | 0.58 | (0.27–1.27) | 0.277 | base |
| **hosp** | High Kyna / Balanced | 0.68 | (0.38–1.23) | 0.346 | base |
| **hosp** | High Quin / Balanced | 1.18 | (0.58–2.39) | 0.936 | base |
| **procedure** | Low Kyna / High Kyna | 1.9 | (0.86–4.23) | 0.162 | base |
| **procedure** | Low Kyna / High Quin | 1.11 | (0.44–2.79) | 0.992 | base |
| **procedure** | Low Kyna / Balanced | 2.1 | (1.07–4.14) | 0.0246 | base |
| **procedure** | High Kyna / High Quin | 0.58 | (0.22–1.53) | 0.475 | base |
| **procedure** | High Kyna / Balanced | 1.11 | (0.5–2.44) | 0.988 | base |
| **procedure** | High Quin / Balanced | 1.9 | (0.75–4.82) | 0.284 | base |
| **any** | Low Kyna / High Kyna | 1.62 | (1.11–2.35) | 0.0054 | base |
| **any** | Low Kyna / High Quin | 1.11 | (0.71–1.76) | 0.931 | base |
| **any** | Low Kyna / Balanced | 1.39 | (1.02–1.91) | 0.0346 | base |
| **any** | High Kyna / High Quin | 0.69 | (0.44–1.07) | 0.136 | base |
| **any** | High Kyna / Balanced | 0.86 | (0.62–1.2) | 0.651 | base |
| **any** | High Quin / Balanced | 1.25 | (0.82–1.92) | 0.522 | base |
| **biol** | Low Kyna / High Kyna | 1.16 | (0.72–1.88) | 0.856 | full |
| **biol** | Low Kyna / High Quin | 1.11 | (0.6–2.05) | 0.969 | full |
| **biol** | Low Kyna / Balanced | 1.1 | (0.74–1.64) | 0.926 | full |
| **biol** | High Kyna / High Quin | 0.96 | (0.51–1.8) | 0.998 | full |
| **biol** | High Kyna / Balanced | 0.95 | (0.62–1.46) | 0.989 | full |
| **biol** | High Quin / Balanced | 0.99 | (0.55–1.79) | 1 | full |
| **steroid** | Low Kyna / High Kyna | 1.44 | (0.85–2.45) | 0.289 | full |
| **steroid** | Low Kyna / High Quin | 0.81 | (0.44–1.49) | 0.809 | full |
| **steroid** | Low Kyna / Balanced | 0.92 | (0.6–1.39) | 0.948 | full |
| **steroid** | High Kyna / High Quin | 0.56 | (0.29–1.07) | 0.1 | full |
| **steroid** | High Kyna / Balanced | 0.63 | (0.39–1.02) | 0.0696 | full |
| **steroid** | High Quin / Balanced | 1.13 | (0.63–2.04) | 0.947 | full |
| **hosp** | Low Kyna / High Kyna | 1.46 | (0.71–2.98) | 0.531 | full |
| **hosp** | Low Kyna / High Quin | 0.96 | (0.43–2.16) | 0.999 | full |
| **hosp** | Low Kyna / Balanced | 0.98 | (0.56–1.69) | 0.999 | full |
| **hosp** | High Kyna / High Quin | 0.66 | (0.28–1.58) | 0.614 | full |
| **hosp** | High Kyna / Balanced | 0.67 | (0.35–1.28) | 0.388 | full |
| **hosp** | High Quin / Balanced | 1.01 | (0.46–2.22) | 1 | full |
| **procedure** | Low Kyna / High Kyna | 1.26 | (0.54–2.93) | 0.897 | full |
| **procedure** | Low Kyna / High Quin | 0.91 | (0.35–2.39) | 0.995 | full |
| **procedure** | Low Kyna / Balanced | 1.41 | (0.69–2.89) | 0.613 | full |
| **procedure** | High Kyna / High Quin | 0.72 | (0.26–2.02) | 0.85 | full |
| **procedure** | High Kyna / Balanced | 1.12 | (0.5–2.51) | 0.985 | full |
| **procedure** | High Quin / Balanced | 1.54 | (0.59–4.06) | 0.654 | full |
| **any** | Low Kyna / High Kyna | 1.2 | (0.8–1.81) | 0.667 | full |
| **any** | Low Kyna / High Quin | 0.87 | (0.53–1.44) | 0.893 | full |
| **any** | Low Kyna / Balanced | 1.02 | (0.72–1.45) | 0.998 | full |
| **any** | High Kyna / High Quin | 0.73 | (0.44–1.21) | 0.368 | full |
| **any** | High Kyna / Balanced | 0.85 | (0.59–1.23) | 0.675 | full |
| **any** | High Quin / Balanced | 1.17 | (0.73–1.89) | 0.824 | full |

**Supplemental Table 7.** Contingency table of aggressive vs. non-aggressive disease course by metabolomic cluster in the Ibsen III cohort.

| **Metabotype** | **Aggressive** | **Not Aggressive** |
| --- | --- | --- |
| Low Kyna | 8 | 42 |
| High Kyna | 10 | 132 |
| High Quin | 17 | 82 |
| Balanced | 16 | 273 |

**Supplemental Table 8.** Pairwise Fisher’s exact test p-values (unadjusted and BH-adjusted) comparing rates of aggressive disease between metabolomic clusters.

| **Comparison** | **p_raw_** | **p_adj_** |
| --- | --- | --- |
| Low Kyna vs High Kyna | 0.087 | 0.175 |
| Low Kyna vs High Quin | 1.0 | 1.0 |
| Low Kyna vs Balanced | 0.12 | 0.179 |
| High Kyna vs High Quin | 0.022 | 0.098 |
| High Kyna vs Balanced | 0.685 | 0.821 |
| High Quin vs Balanced | 0.033 | 0.098 |

**Supplemental Table 9.** Modelling results for the NAD-related metabolites measured in the MSCCR cohort. Linear models were adjusted for age and sex assigned at birth.

| **Metabolite** | **Contrast** | **Estimate** | **Standard error** | **p-value (raw)** |
| --- | --- | --- | --- | --- |
| **1-methylnicotinamide** | Low Kyna - High Kyna | -0.73 | 0.11 | <0.001 |
| **1-methylnicotinamide** | Low Kyna - High Quin | -0.79 | 0.15 | <0.001 |
| **1-methylnicotinamide** | Low Kyna - Balanced | -0.39 | 0.1 | <0.001 |
| **1-methylnicotinamide** | High Kyna - High Quin | -0.05 | 0.14 | 0.98 |
| **1-methylnicotinamide** | High Kyna - Balanced | 0.34 | 0.1 | 0.0026 |
| **1-methylnicotinamide** | High Quin - Balanced | 0.4 | 0.14 | 0.02 |
| **quinolinate** | Low Kyna - High Kyna | 0.46 | 0.1 | <0.001 |
| **quinolinate** | Low Kyna - High Quin | -0.78 | 0.12 | <0.001 |
| **quinolinate** | Low Kyna - Balanced | 1.05 | 0.08 | <0.002 |
| **quinolinate** | High Kyna - High Quin | -1.24 | 0.12 | <0.003 |
| **quinolinate** | High Kyna - Balanced | 0.59 | 0.08 | <0.004 |
| **quinolinate** | High Quin - Balanced | 1.83 | 0.12 | <0.005 |
| **nicotinamide** | Low Kyna - High Kyna | 0.13 | 0.12 | 0.72 |
| **nicotinamide** | Low Kyna - High Quin | 0.17 | 0.15 | 0.69 |
| **nicotinamide** | Low Kyna - Balanced | -0.08 | 0.1 | 0.88 |
| **nicotinamide** | High Kyna - High Quin | 0.04 | 0.15 | 0.99 |
| **nicotinamide** | High Kyna - Balanced | -0.2 | 0.1 | 0.2 |
| **nicotinamide** | High Quin - Balanced | -0.25 | 0.14 | 0.33 |
| **trigonelline (N'-methylnicotinate)** | Low Kyna - High Kyna | 0 | 0.12 | 1.00 |
| **trigonelline (N'-methylnicotinate)** | Low Kyna - High Quin | -0.07 | 0.15 | 0.97 |
| **trigonelline (N'-methylnicotinate)** | Low Kyna - Balanced | -0.07 | 0.11 | 0.91 |
| **trigonelline (N'-methylnicotinate)** | High Kyna - High Quin | -0.07 | 0.15 | 0.97 |
| **trigonelline (N'-methylnicotinate)** | High Kyna - Balanced | -0.06 | 0.1 | 0.92 |
| **trigonelline (N'-methylnicotinate)** | High Quin - Balanced | 0 | 0.15 | 1.00 |
| **N1-methyl-2-pyridone-5-carboxamide** | Low Kyna - High Kyna | -0.57 | 0.11 | <0.001 |
| **N1-methyl-2-pyridone-5-carboxamide** | Low Kyna - High Quin | -0.99 | 0.14 | <0.001 |
| **N1-methyl-2-pyridone-5-carboxamide** | Low Kyna - Balanced | -0.01 | 0.1 | 1 |
| **N1-methyl-2-pyridone-5-carboxamide** | High Kyna - High Quin | -0.42 | 0.14 | 0.02 |
| **N1-methyl-2-pyridone-5-carboxamide** | High Kyna - Balanced | 0.55 | 0.1 | <0.001 |
| **N1-methyl-2-pyridone-5-carboxamide** | High Quin - Balanced | 0.97 | 0.14 | <0.001 |
